# COVID-19 herd immunity in the Brazilian Amazon

**DOI:** 10.1101/2020.09.16.20194787

**Authors:** Lewis F Buss, Carlos A Prete, Claudia MM Abrahim, Alfredo Mendrone, Tassila Salomon, Cesar de Almeida-Neto, Rafael FO França, Maria C Belotti, Maria PSS Carvalho, Allyson G Costa, Myuki AE Crispim, Suzete C Ferreira, Nelson A Fraiji, Susie Gurzenda, Charles Whittaker, Leonardo T Kamaura, Pedro L Takecian, Marcio K Oikawa, Anna S Nishiya, Vanderson Rocha, Nanci A Salles, Andreza A de Souza-Santos, Martirene A da Silva, Brian Custer, Manoel Barral-Netto, Moritz UG Kraemer, Rafael HM Pereira, Oliver G Pybus, Michael P Busch, Márcia C Castro, Christopher Dye, Vitor H Nascimento, Nuno R Faria, Ester C Sabino

**Author notes:** These authors contributed equally (Lewis F Buss, Carlos A Prete, Claudia MM Abrahim, Alfredo Mendrone, Tassila Salomon, Nuno R Faria, Ester C Sabino).

## Abstract

The herd immunity threshold is the proportion of a population that must be immune to an infectious disease, either by natural infection or vaccination such that, in the absence of additional preventative measures, new cases decline and the effective reproduction number falls below unity^1^. This fundamental epidemiological parameter is still unknown for the recently-emerged COVID-19, and mathematical models have predicted very divergent results^2,3^. Population studies using antibody testing to infer total cumulative infections can provide empirical evidence of the level of population immunity in severely affected areas. Here we show that the transmission of SARS-CoV-2 in Manaus, located in the Brazilian Amazon, increased quickly during March and April and declined more slowly from May to September. In June, one month following the epidemic peak, 44% of the population was seropositive for SARS-CoV-2, equating to a cumulative incidence of 52%, after correcting for the false-negative rate of the antibody test. The seroprevalence fell in July and August due to antibody waning. After correcting for this, we estimate a final epidemic size of 66%. Although non-pharmaceutical interventions, plus a change in population behavior, may have helped to limit SARS-CoV-2 transmission in Manaus, the unusually high infection rate suggests that herd immunity played a significant role in determining the size of the epidemic.

## Main text

There is no consensus on what proportion of a population must be infected with SARS-CoV-2 before herd immunity is reached, the threshold above which each infection leaves less than one secondary infection and new cases decline in the absence of other control measures^1^. Estimates of this threshold can help to inform aspects of public health policy, including decisions to reopen society and the roll-out and impact of vaccination campaigns. Given a basic reproduction number (R_0_) of 2.5^4^, the theoretical herd immunity threshold for SARS-CoV-2 under simple epidemiological models is ~60%. However models that account for heterogenous population mixing predict lower values, ranging from 20%^3^ to 43%^2^. The herd immunity threshold, together with social distancing and other control measures, determine the final epidemic size.

Antibody prevalence studies employ serology testing to measure the proportion of a population with evidence of prior infection. When conducted in a given location, a serial cross-sectional seroprevalence study design can provide empirical evidence of the final epidemic size. Although there have been numerous antibody prevalence studies in Europe and North America, the comparatively low estimates of cumulative infections there (generally <20% ^5–7^) cannot be taken to reflect herd immunity due to the widespread adoption of effective non-pharmaceutical control measures in those locations^8^.

In contrast, Brazil has one of the most rapidly-growing COVID-19 epidemics in the world, with the Amazon (Northern Brazil) being the worst hit region^9^. Manaus is the capital of Amazonas state with a population of over two million and population density of 158 inhabitants/km^2^. The first case in Manaus was confirmed on 13^th^ March 2020^10^ and was followed by an explosive epidemic; excess mortality in Manaus in the first week of May was 4.5 times that of the preceding year ^11^. The epidemic peak in early May was followed by a sustained drop in cases and deaths despite relaxation of control measures (Table S3).

Although the ideal design to determine prevalence of SARS-CoV-2 infection is a population-based sample, this approach is time consuming and expensive. Routine blood donations can serve as a logistically-tractable alternative ^12-14^ Herein, we present cross-sectional monthly seroprevalence estimates in blood donors in Manaus spanning the first seven months of transmission in Brazil and correlate these findings with the entire epidemic curve in the Amazon region. We compare these estimates with parallel findings from São Paulo (southeast Brazil), where the first COVID-19 cases were detected in Brazil ^4,15^

## SARS-CoV-2 antibody dynamics and serology assay validation

We used a commercially available chemiluminescence assay (CIMA) that detects IgG antibody against the SARS-CoV-2 nucleocapsid (N) protein (Abbott, Chicago, USA). To infer the true prevalence of infections from antibody prevalence, the sensitivity and specificity of the antibody test need to be accounted for^16^. The specificity of the Abbott SARS-CoV-2 CIMA has consistently been shown to be high (>99.0%)^7,17,18^. However, the high sensitivity (>90.0%)^7,18^ evidenced in previous validation studies was based on severe COVID-19 cases and may not apply to blood donor screening^19,20^ for two reasons. Firstly, most SARS-CoV-2 infections in blood donors are asymptomatic. The weaker antibody response in asymptomatic disease^21^ may lead to a lower initial seroconversion rate. Second, due to antibody waning, the sensitivity falls over time after infection^22^. As the case mix varies through the course of an epidemic – proportionally more recent cases at the start with increasingly remote cases through time – the sensitivity will drop as a result of seroreversion (transition from a positive to negative assay result).

We used a variety of clinical samples at different time points to gain insight into the dynamics of the anti-N IgG detected by the Abbott CIMA (Fig. 1). In COVID-19 hospitalized patients at 20-33 days post symptom onset, the sensitivity was 91.8% (95% confidence interval, CI, 80.8% to 96.8%), reflecting high disease severity and optimal timing of blood collection, but also suggesting that ~8% of severe convalescent cases do not develop detectable antibodies. Among a cohort of symptomatic cases with mild disease also tested in the early convalescent period, the sensitivity fell to 84.5% (95%CI 78.7% to 88.9%) – indicating initial seroconversion is lower in mild cases. In samples drawn later (50-131 days) from the same cohort, the sensitivity was lower still (80.4%, 95%CI 71.8% to 86.8%), reflecting antibody waning. Indeed, in a subset of 104 patients with two consecutive blood draws, the signal-to-cutoff (S/C) clearly declined over the period observed (Fig. 1B) and among 88 individuals with a positive reading at the first time point, the mean rate of decay was -0.9 log2 S/C units every 100 days (95%CI -1.1 to -0.75), equating to a half-life of 106 days (95%CI 89 to 132 days) (Fig. 1C).

Finally, we tested 1,000 blood donations given in São Paulo in July 2020 in parallel using a second high-specificity (>99.0%^23^) immunoassay (Roche Elecsys, Rotkreuz, Switzerland). One-hundred and three samples were positive on the Abbott CIMA and an additional 30 were positive on the second assay. Assuming all 133 samples were true positives the sensitivity of the Abbott N IgG assay was 77.4% (95%CI 69.6% to 83.7%) on asymptomatic blood donor serosurveillance samples. The Roche assay detects total Ig and the signal is more stable than the Abbott assay^22^. As samples in July were donated four months into the on-going epidemic in São Paulo, the false negatives on the Abbott assay include both cases that did not initially seroconvert (“serosilent” infections), as well as remote infections with subsequent seroreversion.

Because the specificity was high, with only one false-positive result in 821 pre-epidemic donations from Manaus, we also attempted to improve assay performance by reducing the threshold for a positive result from 1.4 S/C (as per the manufacturer) to 0.4 S/C. The 27 false-positives resulted in a specificity of 96.7%. The sensitivities at this threshold are shown in Table S1.

**Fig. 1.**
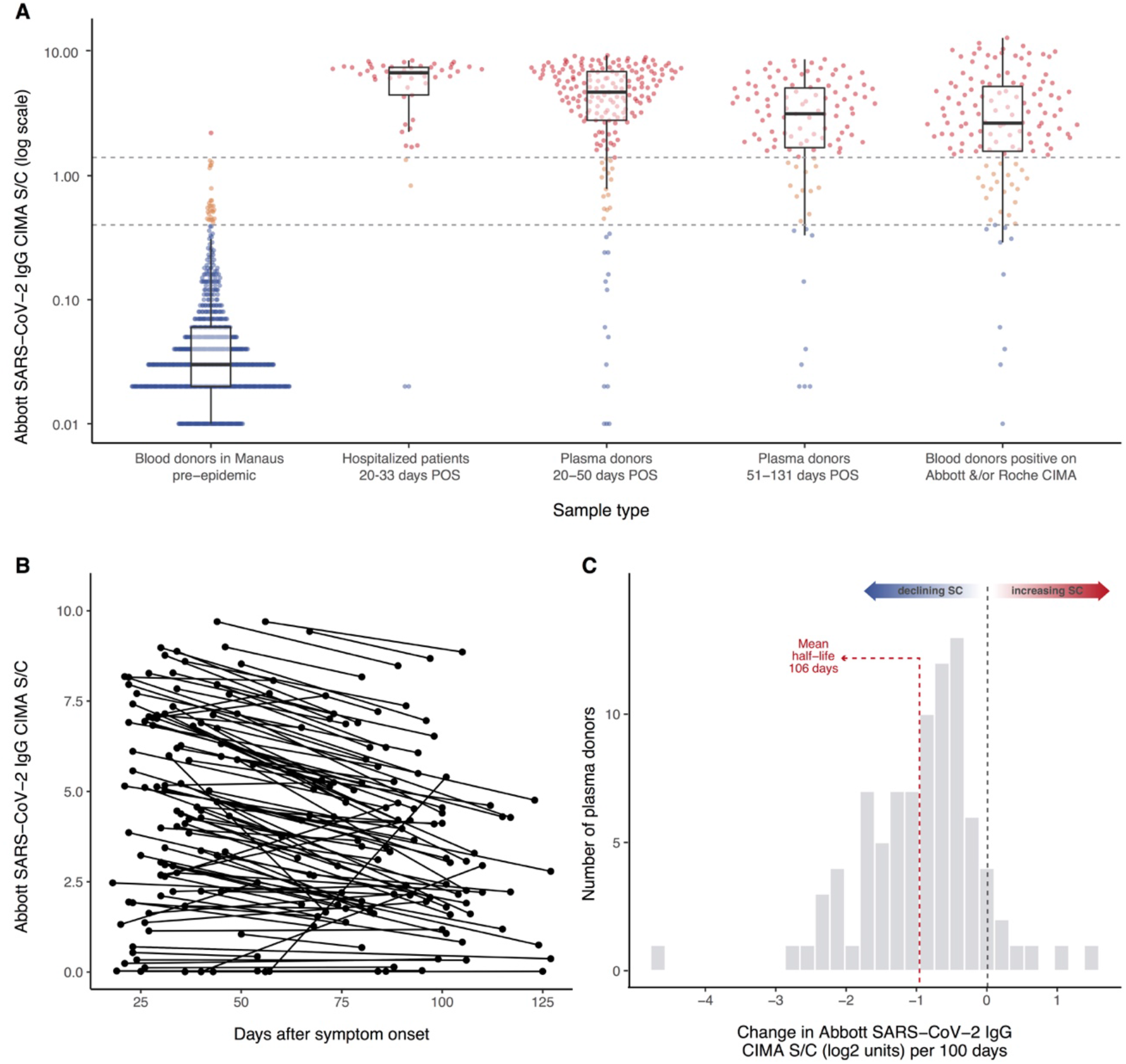
Abbott SARS-CoV-2 N IgG chemiluminescence assay performance and antibody dynamics in different clinical samples. Panel A – signal-to-cutoff (S/C) values on the Abbott chemiluminescence assay (CIMA) in the following clinical samples (from left to right): 821 routine blood donation samples made in Manaus in February 2020, more than 1 month prior to the first case notified in the city; 49 samples collected at 20-33 days after symptom onset from SARS-CoV-2-PCR positive patients requiring hospital care; 193 patients with PCR-confirmed symptomatic COVID-19 not requiring hospital care, with plasma donation samples taken in the early convalescent period; 107 samples from the same non-hospitalized plasma donor cohort from the late convalescent period; 133 samples that tested positive on either the Abbott CIMA or the Roche Elecsys assay out of 1,000 routine blood donations collected in July 2020 and tested in parallel from the Fundação Pró-Sangue blood center (São Paulo). Upper dashed line - manufacturer’s threshold for positive result of 1.4 S/C; lower dashed line - alternative threshold of 0.4 S/C. Panel B - 104 convalescent plasma donors with two blood draws for serology testing on the Abbott CIMA. Panel C - histogram of the slopes among 88 individuals shown in panel B that tested positive (>1.4 S/C) at the first time point. POS = post onset of symptoms.

### Prevalence of SARS-CoV-2 antibodies in Manaus and São Paulo

In order to estimate the proportion of the population with antibodies against SARS-CoV-2, we used a convenience sample of routine blood donations made at the Fundação Pró-Sangue blood bank in São Paulo and the Fundação Hospitalar de Hematologia e Hemoterapia do Amazonas (HEMOAM) in Manaus. The monthly sample size and sampling dates, spanning February to August, are shown in Table 1.

**Table 1.** Results of cross-sectional samples of blood donors in Manaus and São Paulo. Weighted prevalence was calculated by applying weights proportional to the projected age-sex population structure of Manaus and São Paulo within the age group eligible to donate blood. Further adjustment for sensitivity and specificity was performed with the Rogan and Gladen method ^24^ to give the adjusted prevalence at each time point (i.e. sensitivity/specificity adjustment was in addition to age-sex re-weighting). At the 1.4 S/C threshold the sensitivity and specificity were taken to be 84.0% and 99.9%, respectively; at the 0.4 threshold they were taken to be 92.2% and 96.7%, respectively (see Table S1). See Methods for details of the seroreversion correction.

| Location and sampling dates | Total samples tested | 1.4 S/C threshold to define positive result |  |  |  |  | 0.4 S/C threshold to define positive result |  |  |  |
| --- | --- | --- | --- | --- | --- | --- | --- | --- | --- | --- |
|  |  | Positive samples | Crude prevalence % (95% CI) | Weighted prevalence (95% CI) | Sensitivity and specificity adjusted prevalence (95% CI) | Seroreversion adjusted prevalence (95%CI) | Positive samples | Crude prevalence % (95% CI) | Weighted prevalence % (95% CI) | Sensitivity and specificity adjusted prevalence % (95% CI) |
| **Manaus** |  |  |  |  |  |  |  |  |  |  |
| Feb 7th-13th | 821 | 1 | 0.1 (0.0-0.7) | 0.4 (0.0-2.2) | 0.3 (0.0-2.4) | - | 27 | 3.3 (2.2-4.7) | 3.7 (2.0-6.3) | 0.4 (0.0-3.3) |
| Mar 6th-12th | 831 | 6 | 0.7 (0.3-1.6) | 0.7 (0.2-1.8) | 0.7 (0.1-2.0) | 0.7 (0.2-1.6) | 25 | 3.0 (2.0- 4.4) | 2.6 (1.6- 4.1) | 0.0 (0.0-0.9) |
| Apr 6th-17th | 829 | 46 | 5.5 (4.1- 7.3) | 4.1 (2.8-5.8) | 4.8 (3.3-6.8) | 5.0 (3.7-6.6) | 84 | 10.1 (8.2-12.4) | 7.7 (5.9-9.9) | 5.0 (2.9-7.4) |
| May 5th-14th | 900 | 359 | 39.9 (36.7-43.2) | 37.4 (33.0-42.0) | 44.2 (39.0-49.7) | 45.9 (41.7-50.6) | 413 | 45.9 (42.6-49.2) | 42.1 (37.6-46.7) | 43.6 (38.5-48.8) |
| Jun 5th-15th | 909 | 421 | 46.3 (43.0-49.6) | 43.8 (39.6-48.0) | 51.8 (46.8-56.8) | 64.8 (59.7-74.6) | 494 | 54.3 (51.0-57.6) | 52.5 (48.2-56.7) | 55.3 (50.5-60.1) |
| Jul 6th-15th | 1145 | 418 | 36.5 (33.7-39.4) | 33.9 (30.0-37.9) | 40.0 (35.5-44.8) | 66.1 (60.8-79.8) | 580 | 50.7 (47.7-53.6) | 46.9 (42.7-51.1) | 49.0 (44.3-53.8) |
| Aug 8th-19th | 881 | 242 | 27.5 (24.5-30.5) | 25.5 (22.2-29.1) | 30.1 (26.2-34.3) | 66.1 (60.8-79.9) | 426 | 48.4 (45.0-51.7) | 44.0 (40.0-48.0) | 45.7 (41.3-50.2) |
| **São Paulo** |  |  |  |  |  |  |  |  |  |  |
| Feb 8th-29th | 799 | 7 | 0.9 (0.4-1.8) | 0.9 (0.3-1.9) | 1.0 (0.3-2.1) | - | 36 | 4.5 (3.2-6.2) | 4.2 (2.9-6.0) | 1.0 (0.0-2.9) |
| Mar 9th-21st | 2454 | 22 | 0.9 (0.6-1.4) | 0.8 (0.5-1.2) | 0.8 (0.5-1.3) | 0.8 (0.5-1.2) | 149 | 6.1 (5.2-7.1) | 5.8 (4.9-6.8) | 2.8 (1.8-3.9) |
| Apr 8th-30th | 900 | 27 | 3.0 (2.0-4.3) | 2.6 (1.6-3.9) | 2.9 (1.7-4.5) | 3.1 (2.2-4.4) | 58 | 6.4 (4.9-8.3) | 6.8 (4.9-9.1) | 4.0 (1.8-6.6) |
| May 8th-21st | 826 | 44 | 5.3 (3.9-7.1) | 5.1 (3.4-7.2) | 5.9 (4.3-9.2) | 6.9 (5.4-9.1) | 69 | 8.4 (6.6-10.5) | 7.5 (5.6-9.8) | 4.8 (2.6-7.4) |
| Jun 8th-20th | 880 | 105 | 11.9 (9.9-14.3) | 11.6 (9.3-14.1) | 13.6 (12.0-18.1) | 16.1 (14.0-19.4) | 145 | 15.3 (13.0-17.9) | 14.9 (12.5-17.8) | 13.2 (10.3-16.3) |
| Jul 13th-25th | 879 | 84 | 9.6 (7.7-11.7) | 9.5 (7.6-11.8) | 11.2 (8.8-13.9) | 17.2 (15.5-21.0) | 116 | 13.2 (11.1-15.6) | 12.8 (10.5-15.3) | 10.7 (8.1-13.5) |
| Aug 10th-21st | 813 | 98 | 12.1 (9.9- 13.3) | 11.6 (9.2-14.3) | 13.6 (10.8 -16.8) | 22.4 (19.9-27.6) | 149 | 18.3 (15.7-21.2) | 16.7 (13.9-19.6) | 15.1 (12.0-18.5) |

Table 1 also presents the crude monthly antibody prevalence among blood donors; the prevalence re-weighted to the age-sex distribution of each city; and the prevalence following adjustment for test performance, calculated both at the manufacture’s threshold (1.4 S/C) and the reduced threshold (0.4 S/C) (see above). Sensitivity adjustments were based on the early-phase convalescent plasma donors (Fig. 1A), as these estimates account for initial non-seroconversion before significant antibody waning. We then account for antibody waning using a simple model-based approach (see Methods and Table 1). Antibody prevalence according to demographic categories is shown in Table S2.

The prevalence of SARS-CoV-2 antibodies in February and March was low (<1%) in both São Paulo and Manaus. This is consistent with the timing of the first confirmed cases that were diagnosed on 13^th^ March in Manaus, and on the 25^th^ of February in São Paulo^10^.

In Manaus, after adjustment for the sensitivity and specificity of the test, and re-weighting for age and sex, the prevalence of SARS-CoV-2 IgG antibodies was 4.8% (95%CI 3.3%-6.8%) in April, 44.2% (95%CI 39.0%-49.7%) in May, reaching a peak of 51.8% (46.8%-56.8%) in June (Fig. 2). The increasing seroprevalence closely followed the curve of cumulative deaths. In São Paulo the prevalence of SARS-CoV-2 IgG in blood donors also increased steadily, reaching 13.6% (12.0%-18.1%) in June.

**Fig. 2.**
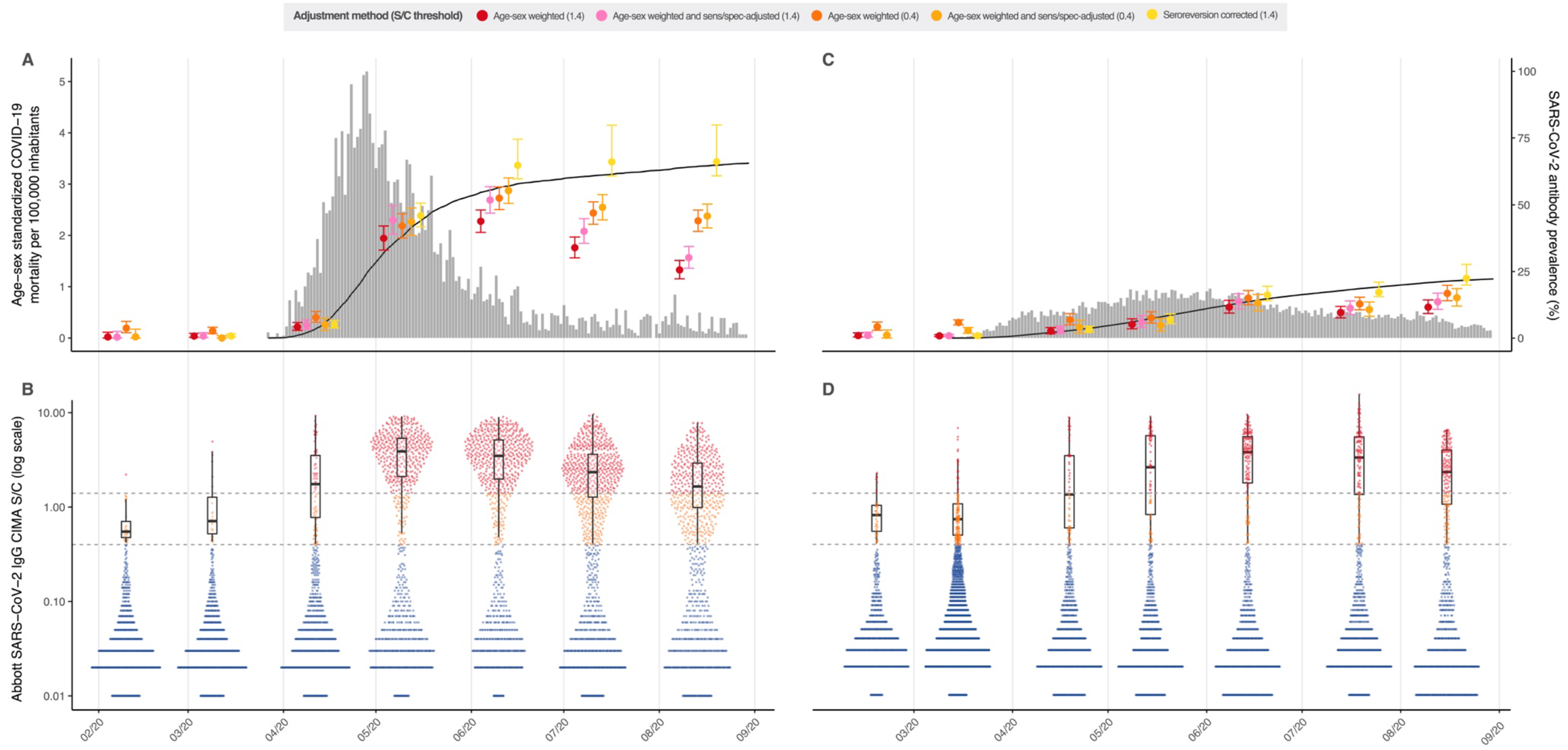
Monthly antibody prevalence and signal-to-cutoff (S/C) reading in Manaus and São Paulo. SARS-CoV-2 antibody prevalence estimates in Manaus (A) and São Paulo (C) with a range of corrections. Error bars are 95% confidence intervals. Grey bars are standardized daily mortality using confirmed COVID-19 deaths in the SIVEP-Gripe (https://covid.saude.gov.br/) notification system and standardized by the direct method using the total projected Brazilian population for 2020 as the reference. Black lines are the cumulative deaths rescaled so that the maximum is set to the maximum seroprevalence estimate for each city. Mortality data is plotted according to the date of death. Distribution of S/C values over the seven monthly samples are shown for Manaus (B) and São Paulo (D). Each point represents the S/C reading for a single donation sample. Upper dashed line - manufacturer’s threshold (1.4 S/C units); lower dashed line - alternative threshold (0.4 S/C units); black boxplots show the median, interquartile range and range of S/C values above 0.4 (i.e., excluding very low and likely true-negative values.

Between June and August, the effect of seroreversion became apparent in both cities. In Manaus, following the peak antibody prevalence in June, the proportion of blood donors testing positive fell to 40.0% in July, and 30.1% in August. Excluding extreme negative samples (<0.4 S/C), the median assay signal fell steadily from May onwards: 3.9 (May), 3.5 (June), 2.3 (July) and 1.7 (August), see Fig. 2B. Similarly, in São Paulo the antibody prevalence remained stable between June and August, while the number of daily COVID-19 deaths also remained relatively stable, reflecting the balance between antibody waning from infection earlier in the outbreak and seroconversion following recent infections (Fig. 2C).

In Manaus, the effect of antibody waning on apparent prevalence was partially ameliorated by reducing the threshold for a positive result from 1.4 S/C to 0.4 S/C and correcting for the resulting increased false-positive rate. However, the results in São Paulo were largely unchanged by this correction (Fig. 2 and Table 1).

We further correct for seroreversion with a model-based approach (see Methods for details). Briefly, we assume that the probability of remaining seropositive decays exponentially from the time of recovery. We estimate the decay rate and the proportion of patients that serorevert using the seroprevalence data from Manaus to find the minimum decay rate that minimizes the number of new cases in July and August while avoiding drops in prevalence – i.e. assuming there were few cases in Manaus in July and August and changes in seroprevalence were due mainly to waning antibodies. The results of these corrections are shown in Table 1 and Fig. 2. We find that after adjusting for seroreversion, the cumulative incidence of infections in Manaus may have reached as high as 66.1% (95%CI 60.8%-79.9%). Although this is the minimum prevalence estimate allowed by the exponential decay model, and should therefore be conservative, in the absence of an accepted approach to account for seroreversion, these results should be interpreted with caution. The reliability of this estimate depends on the validity of the exponential decay assumption.

### Infection fatality ratio in Manaus

In Manaus the overall fatality ratio (IFR) was 0.17% and 0.28%, considering PCR confirmed COVID-19 deaths and probable COVID-19 deaths based on syndromic identification, respectively; whereas in São Paulo, the global IFRs were 0.46% and 0.72%, respectively. The difference may be explained by an older population structure in São Paulo (Fig. S1). Supporting this inference, the age-specific IFRs were similar in the two cities, and similar to estimates based on data from Wuhan, China^25^ (Fig. S1B).

## Discussion

Our results show that between 44% and 66% of the population of Manaus was infected with SARS-CoV-2 through the course of the epidemic. The lower estimate does not account for false-negative cases or antibody waning; the upper estimate accounts for both. The elevated mortality and the rapid and sustained drop in cases (Figure 2A and S4) suggest population immunity played a significant role in determining the size of the epidemic in Manaus.

The non-pharmaceutical interventions implemented in the city of Manaus (Table S3) were similar to other cities in Brazil including São Paulo. They were implemented in late March before the epidemic took off. Furthermore, cell phone mobility data showed a marked increase in physical distancing beginning in mid-March, with a similar pattern over time to São Paulo (Fig. S2). Therefore, it remains unclear what accounted for such rapid transmission of SARS-CoV-2 in Manaus. Possible explanations include low socioeconomic conditions, with household crowding^26^, limited access to clean water, and reliance on high-risk boat travel,^9^ in which over-crowding results in accelerated contagion, similar to that seen on cruise ships ^27^ The young mobile population with potentially low pre-existing immunity to SARS-CoV-2^28^, as well as the circulation of multiple virus lineages introduced from multiple locations^10^ may have contributed to the large scale of the outbreak.

Our results cannot be extrapolated directly to other contexts due to differences in population demographics, behavior, vulnerability to infection, as well as implementation and adherence to non-pharmaceutical measures. The proportion of the population with immunity to SARS-CoV-2 works in tandem with these factors to tip the effective reproduction number below unity. Indeed, given a basic reproduction number (R_0_) of 2.5 (as estimated for Amazonas state^4^ and Manaus - see Fig. S3) the herd immunity threshold in Manaus would be 60%, and the final size of the epidemic 89%. This assumes that the population was mixing homogeneously and not subject to effective NPIs. Homogeneous mixing is unlikely to be a valid assumption;^2^ and heterogeneous exposure or susceptibility to infection may explain why the estimated final size of 44-66% infected is less than 89%.

We observed a waning of antibodies following the epidemic peak in Manaus. These findings have significant implications for the design and interpretation of antibody prevalence studies. For the purpose of estimating total cumulative infections in a population, the assay chosen should ideally detect a long-lasting component of the humoral response to SARS-CoV-2. Although other assays such as the Roche and Ortho Total Ig assays seem promising in this regard, caution is required before extrapolating data from symptomatic patient cohorts ^22^ to population surveys, as most infections are asymptomatic in this use case. Despite this limitation of the Abbott assay, one potential advantage to the decay in signal over time is to monitor for reinfections at the population level in the case of a second epidemic wave based on boosting of seroreactivity. Indeed, Manaus may act as a sentinel to determine the longevity of population immunity and frequency of reinfections. An additional strategy to antibody surveillance would be monitoring of local versus imported cases, with a relative increase in local cases suggesting population immunity was no longer preventing onwards transmission.

Another important limitation is the extent to which blood donors are representative of the wider population with respect to SARS-CoV-2 exposure. Firstly, children and the elderly are excluded from blood donation. The eligible age range for blood donation in Brazil (16 - 69yr), as well as sex distributions in donors, are different from the underlying populations in both cities (Fig. S4); however, we attempted to account for this by re-weighting according to age and sex. Furthermore, only healthy asymptomatic adults without a recent history of COVID-19 infection are eligible to donate blood. This would be expected to lead to an underestimation of true prevalence – the healthy volunteer donor effect. It is reassuring that a household survey in São Paulo city, employing a random sampling strategy and comparable antibody assay, found very similar results to our study: 4.7% seroprevalence in May ^29^ (versus 5.3% in blood donors) and 11.4% in June (versus 11.9% in blood donors), and that the age-specific IFRs for both São Paulo and Manaus were similar to those estimated for China using different methods (Fig S1B)^25^

Finally, in another population-based serosurvey conducted in mid-May in Manaus^9^, the SARS-CoV-2 seroprevalence was found to be 12.5%, less than half the prevalence at this time point (5th to 14th) among blood donors. This discrepancy is likely accounted for by the lower sensitivity of the assay that was used to test capillary (finger prick) blood. Although the authors corrected for test characteristics, it is likely that the true sensitivity in capillary blood is lower ^30^. This highlights the advantage of using the blood donor population, where the infrastructure necessary for the use of state-of-the-art laboratory-based serological assays on blood samples is well established. Furthermore, blood donors may enable longitudinal prospective monitoring of infections, immune persistence and rates of reinfections, and facilitate surveillance in areas of the globe where population studies are too expensive to maintain.

## Data Availability

The data required to reproduce the results in this article will be deposited on the Figshare repository upon acceptance of the article (URL), where the raw data underlying the main figures will be provided. Also, upon acceptance, the custom code will be made available at the linked GitHub repository (URL). 

## Methods

### Ethics

This project was approved by the Brazilian national research ethics committee, CONEP CAAE - 30178220.3.1001.0068.

### Study sites and setting

This report is part of a wider study (Covid-IgG) monitoring SARS-CoV-2 antibody prevalence among blood donors in eight Brazilian cities (Belo Horizonte, Curitiba, Fortaleza, Recife, Rio de Janeiro, Salvador, São Paulo and Manaus). The results of this preliminary report are from two participating blood banks: the Fundação Pró-Sangue (FPS) in São Paulo and the Fundação Hospitalar de Hematologia e Hemoterapia do Amazonas (HEMOAM) in Manaus.

### Selection of blood samples for serology testing

Both the FPS and HEMOAM blood centers routinely store residual blood samples for six months after donation. In order to cover a period starting from the introduction of SARS-CoV-2 in both cities, we retrieved stored samples covering the months of February to May in São Paulo, and February to June in Manaus, at which point testing capacity became available. In subsequent months blood samples were prospectively selected for testing. The monthly target was to test 1,000 samples at each study site. However, due to problems with purchasing the kits, supply chain issues, and the period of test validity, some months were under and others over the target (to avoid wasting kits soon to expire). We aimed to include donations starting from the second week of each month (see Table 1 for exact sampling windows).

Part of the remit of the wider project is to develop a system to prospectively select blood donation samples, based on the donor’s residential address, so as to capture a spatially representative sample of each participating city. For example, FPS receives blood donations from people living across the whole greater metropolitan region of São Paulo. The spatial distribution of donors does not follow the population density, with some areas over- and others under-represented. We used residential zip codes (recorded routinely at FPS) to select only individuals living within the city of São Paulo. We then further divided the city into 32 regions (*subprefeituras*) and used their projected population sizes for 2020 to define sampling weights, such that the number of donors selected in any given *subprefeitura* was proportional to the population size. We piloted this approach in São Paulo and have developed an information system to operationalize this process at the participating center. However, at the time of data collection the system was not implemented in HEMOAM and therefore it was not possible to use this sampling strategy. As such, we simply tested consecutive blood donations, beginning from the second week of each month until the target was reached. The spatial distribution of blood donors tested in the study is shown in Fig. S6.

### Abbott SARS-CoV-2 IgG chemiluminescence microparticle assay

We used the Abbott SARS-CoV-2 chemiluminescence microparticle assay (CMIA) that detects IgG antibody against SARS-CoV-2 nucleocapsid protein. The chemiluminescence reaction is measured in relative light units (RLU) that increase as a function of the amount of anti-SARS-CoV-2 IgG antibodies present in the sample. Readings are expressed as the ratio (denoted S/C) between the RLU produced by the sample and the RLU from the system calibrator.

### In-house validation of the Abbott CMIA

Although the Abbott CIMA has been validated in a number of studies ^1–3^ with high specificity (>99.0%) and sensitivity (generally 85-100%), the test characteristics - particularly sensitivity - are expected to vary with the use case and population in which the test is applied. Most validation studies suffer from spectrum bias, enrolling primarily moderate to severe cases as the positive controls to define sensitivity. This will bias estimates of sensitivity upwards, thus causing an underestimation of cumulative infections after correction for test characteristics.

To address this issue, we performed a local validation of the Abbott CIMA on a range of clinical samples. Firstly, we tested samples collected from hospitalized patients with PCR-confirmed SARS-CoV-2 infection at two hospitals in São Paulo (*Hospital das Clínicas* and *Hospital Sírio-Libanês*). All samples were collected at least 20 days after symptom onset. Second, we tested a cohort of volunteer convalescent plasma donors that had milder disease, not requiring historical admission. Samples were collected at two time points following symptom onset: first in the early convalescent period, and second at > 2 months POS. Finally, we tested 1000 routine blood donation samples at the FPS from July 2020 using the Abbott assay and the Roche Elecsys SARS-CoV-2 electro chemiluminescence assay (ECIMA). In July, the pre-test probability of prior SARS-CoV-2 infection in São Paulo was high (>12%) and the Roche ECIMA has a high (>99%) specificity. Therefore, we assumed that any sample that was positive on at least one test to be a true-positive.

### Quantifying antibody waning and rate of seroreversion

We sought to quantify the rate of decline of the anti-nucleocapsid IgG antibody that is detected by the Abbott CMIA. We tested paired serum samples from our cohort of convalescent plasma donors (described above). We calculated the rate of signal decay as the difference in log2 S/C between the first and second time points divided by the number of days between the two visits. We used simple linear regression to determine the mean slope and 95% CI.

### Analysis of seroprevalence data

Using the manufacturer’s threshold of 1.4 S/C to define a positive result we first calculated the monthly crude prevalence of anti-SARS-CoV-2 antibodies as the number of positive samples/total samples tested. The 95% confidence intervals (CI) were calculated by the exact binomial method. We then re-weighted the estimates for age and sex to account for the different demographic make-up of blood donors compared to the underlying populations of São Paulo and Manaus (Fig. S4). Because only people aged between 16 and 70 years are eligible to donate blood, the re-weighting was based on the projected populations in the two cities in this age range only. The population projections for 2020 are available from (https://demografiaufrn.net/laboratorios/lepp/). We further adjusted these estimates for the sensitivity and specificity of the assay using the Rogan and Gladen method^4,5^.

As a sensitivity analysis, we took two approaches to account for the effect of seroreversion through time. Firstly, the manufacturer’s threshold of 1.4 optimizes specificity but misses many true-cases in which the S/C level is in the range of 0.4 – 1.4 (see ref and main text). In addition, individuals with waning antibody levels would be expected to fall initially into this range. Therefore, we present the results using an alternative threshold of 0.4 to define a positive result and adjust for the resultant loss in specificity.

Secondly, we corrected the prevalence with a model-based method assuming that the probability of seroreversion for a given patient decays exponentially with time. We assume that the probability of a recovered individual seroreverting *m* months after recovery is

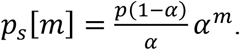

*α* ∊ [0,1] is the monthly attenuation and *p* is the proportion of individuals that can serorevert. The normalization constant

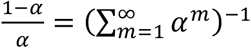

forces

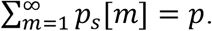

The parameters *α* and *p* are learned using the measured prevalence in Manaus assuming that there are no new recoveries in July and August.

If we denote *R* [*n*] as the cumulative number of recoveries per capita at month *n*, and *S*[*n*] as the cumulative number of seroreversions per capita, then

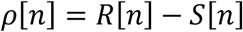

is the measured prevelance,

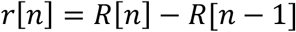

is the number of new recoveries per capita and

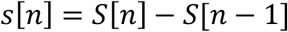

is the number of new seroreversions. Since each recovery at instant *k* contributes on average to 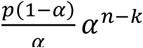 seroreversions at instant *n*, we can model *s*[*n*] as

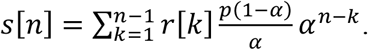

First, we show how to use this equation to estimate *r*[*n*] for fixed parameters (*α*, *p*), and then we show how these parameters are estimated. We define *M* as the number of months with prevalence measurements, the vectors

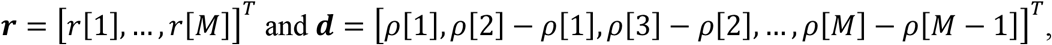

and the *M* × *M* matrix ***A*** whose elements are

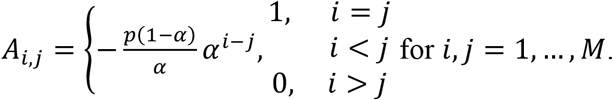

From the measure prevalence *ρ*[*n*], we have

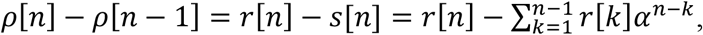

which can be written in matrix form as ***d*** = ***Ar***. As ***A*** is a triangular matrix with ones in the diagonal, it is always invertible, thus ***r*** *=* ***A***−^1^***d***.

In order to estimate *α* and *p*, we generate all pairs of parameters in the set {0.01, 0.02,…,0.99} and compute ***r*** for each (*α,p*) using the prevalence data from Manaus. Since Manaus presents few confirmed cases and deaths in July and August, we estimate (*a^Manaus^, p^Manaus^*) as the parameters that minimize the number of new recoveries in July and August through the minimization of the cost function

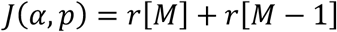

under the constraint *r*[*n*] ≥ 0 for all *n*. These parameters are used to obtain the corrected prevalence in Manaus, which is the cumulative number of recoveries per capita, *R*[*n*]. The same parameters are used to correct the prevalence for São Paulo if they yield a non-negative *r*[*n*] for São Paulo, otherwise they are chosen as the closest parameters to Manaus that produce non-negative *r*[*n*] by minimizing the cost function

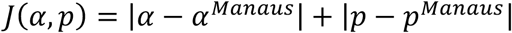

under the constraint *r*[*n*] ≥ 0 for all *n*.

The estimated parameters and their 95% confidence interval for Manaus and São Paulo are: *α^Manaus^ = α^SãoPaulo^* = 0.7352 [0.3236, 0.7744] and *p^Manaus^ = p^SãoPaulo^* = 0.9606 [0.5784, 0.9900]. The estimates and confidence intervals for São Paulo coincide with Manaus.

In the model-based method for correcting the prevalence, only the months between March and August were considered. The measured prevalence used as input for this method was obtained using the manufacturer’s threshold of 1.4, and the correction based on the test specificity (99.9%) and sensitivity (84%) was applied, as well as the normalization by age and sex. Confidence intervals were calculated through bootstrapping, assuming a beta distribution for the input measured prevalence. It is worth noting that even though this model is limited by the exponential decay assumption, assuming distributions with more degrees of freedom may lead to overfitting due to the small number of samples of *ρ*[*n*]. Finally, the obtained values for *α* and *p* must be interpreted as parameters for this model, and not estimates for the actual decay rate and seroreversion probability as they may absorb the effect of variables that are not taken into account by this model.

### Infection fatality ratio

We calculated the global infection fatality ratio in Manaus and São Paulo. The total number of infections was estimated as the product of the population size in each city and the antibody prevalence in June (re-weighted and adjusted for sensitivity and specificity). The number of deaths were taken from the SIVEP-Gripe system, and we used both confirmed COVID-19 deaths, and deaths due to severe acute respiratory syndrome of unknown cause. The latter category likely represents COVID-19 cases in which access to diagnostic testing was limited, and more closely approximate the excess mortality (Fig. S5). We calculated age-specific infection fatality ratios by assuming equal prevalence across all age groups.

### Effective reproduction number

We calculated the effective reproduction number for São Paulo and Manaus using the renewal method^9^, with the serial interval as estimated by Ferguson (2020)^10^. Calculations were made using daily severe acute respiratory syndrome cases with PCR-confirmed COVID-19 in the SIVEP-Gripe system. Region-specific delays between the PCR result release and the date of symptom onset were accounted for using the technique proposed by Lawless (1994)^11^.

### Funding

This study was supported by the Itau Unibanco *Todos pela Saúde* program and CADDE/FAPESP (MR/S0195/1 and FAPESP 18/14389-0). NRF is supported by the Wellcome Trust and Royal Society Sir Henry Dale Fellowship (204311/Z/16/Z). We acknowledge the National Heart, Lung, and Blood Institute of the US National Institutes of Health Recipient Epidemiology and Donor Evaluation Study (REDS, now in its 4^th^ phase, REDS-IV-P) for providing the blood donor demographic and zip code data for analysis (grant number HHSN268201100007I). This work received funding from the U.K. Medical Research Council under a concordat with the U.K. Department for International Development. We additionally acknowledge support from Community Jameel and the NIHR Health Protection Research Unit in Modelling Methodology.

### Authors contributions

Conception - MBN, LFB, MC, BC, CAN, NRF, SCF, AMJ, ASN, RHMP, VR, ECS, NAS, TS, MAS and CW. Acquisition – ACMM, MPSSC, AGC, MAEC, CAN, AASS, NRF, SCF, NAF, PLT, AMJ, MKO, NV, RHMP, VR, ECS, NAS, TS and MAS. Analysis – LFB, CAN, RHMP, CW, ECS, CAP and MCB. Interpretation – ACMM, LFB, MPSSC, AGC, MAEC, CAN, NRF, NAF, ECS, MAS, CW, CD, MUGK, OP; drafting – LFB, ECS; revising – all authors; funding – MBN, AGC, BC, NRF, NAF, ECS, NAS.

### Materials & Correspondence

Correspondence should be directed to Ester C Sabino or Nuno R Faria

### Data availability

The data required to reproduce the results in this article will be deposited on the Figshare repository upon acceptance of the article (URL), where the raw data underlying the main figures will be provided. Also, upon acceptance, the custom code will be made available at the linked GitHub repository (URL).Supplemental Material

**Table S1.** Performance of Abbott SARS-CoV-2 IgG chemiluminescence assay in different clinical samples. The signal-to-cutoff (S/C) of 1.4 is recommended by the manufacturer; 0.4 S/C is a less stringent alternative threshold included as a sensitivity analysis. * Positive samples were identified by testing 1,000 routine donations in parallel on the Abbott CIMA and a second assay (Roche Elecsys IgG ECIMA); positive results on either assay were assumed to be true positives as both have a high specificity.

| S/C threshold and assay result | Negative controls | Positive controls |  |  |  |
| --- | --- | --- | --- | --- | --- |
|  | Blood donations to HEMOAM in Feb<br>n = 821 | Hospitalized patients<br>n = 49 | Plasma donors 20-50d POS<br>n = 193 | Plasma donors 51-131d POS<br>n = 107 | Positive donations to FPS in July *<br>n = 133 |
| **Threshold 1.4 S/C** |  |  |  |  |  |
| Positive | 1 (0.1) | 45 (91.8) | 163 (84.5) | 86 (80.4) | 103 (77.4) |
| Negative | 820 (99.9) | 3 (8.2) | 30 (15.5) | 21 (19.6) | 30 (22.6) |
| **Threshold 0.4 S/C** |  |  |  |  |  |
| Positive | 27 (3.3) | 47 (95.9) | 178 (92.2) | 98 (91.6) | 123 (92.5) |
| Negative | 794 (96.7) | 2 (4.1) | 15 (7.8) | 9 (8.4) | 10 (7.5) |

**Table S2.** Prevalence of SARS-CoV-2 antibodies according to demographic group pooling data for May through August. Odds ratios and 95% confidence intervals (CI) calculated by univariable logistic regression with the reference category denoted by “ref’. Missing data: ethnicity 39 for Manaus and 7 for São Paulo; education - not collected for Manaus, 7 for São Paulo.

|  | Manaus (May-August) |  |  | São Paulo (May-August) |  |  |
| --- | --- | --- | --- | --- | --- | --- |
|  | Negative (<1.4 S/C)<br>n (%) | Positive (≥1.4 S/C)<br>n(%) | OR (95%CI) | Negative (<1.4 S/C)<br>n(%) | Positive (≥1.4 S/C)<br>n(%) | OR (95%CI) |
| **Age (years)** |  |  |  |  |  |  |
| <30 | 1110 (63.1) | 649 (36.9) | 1.0 (ref) | 1137 (90.5) | 119 (9.5) | 1.0 (ref) |
| 30-39 | 633 (60.6) | 412 (39.4) | 1.1 (1.0-1.3) | 845 (91.1) | 83 (8.9) | 0.9 (0.7-1.3) |
| 40-49 | 422 (61.8) | 273 (38.2) | 1.1 (0.9-1.3) | 632 (87.8) | 88 (12.2) | 1.3 (1.0-1.8) |
| 50-59 | 193 (68.2) | 90 (31.8) | 0.8 (0.6-1.0) | 373 (91.2) | 36 (8.8) | 0.9 (0.6-1.4) |
| 60+ | 18 (52.9) | 16 (47.1) | 1.5 (0.8-3.0) | 80 (94.1) | 5 (5.9) | 0.6 (0.2-1.4) |
| **Sex** |  |  |  |  |  |  |
| Female | 792 (69.5) | 347 (30.5) | 1.0 (ref) | 1543 (91.8) | 137 (8.2) | 1.0 |
| Male | 1604 (59.5) | 1093 (40.5) | 1.6 (1.3-1.8) | 1524 (88.7) | 194 (11.3) | 1.4 (1.1-1.8) |
| **Ethnicity** |  |  |  |  |  |  |
| Asian | 17 (68.0) | 8 (32.0) | 0.7 (0.3-1.6) | 75 (94.9) | 4 (5.1) | 0.4 (0.1-0.9) |
| White | 262 (75.9) | 83 (24.0) | 0.5 (0.4-0.6) | 2032 (91.7) | 183 (8.3) | 0.6 (0.5-0.8) |
| Black | 81(67.5) | 39 (32.5) | 0.7(0.5-1.1) | 168 (84.4) | 31 (15.6) | 1.3 (0.8-2.0) |
| Mixed (Pardo) | 2002 (60.8) | 1293 (39.2) | 1.0 (ref) | 784 (87.5) | 112 (12.5) | 1.0 (ref) |
| Indigenous Brazilian | 8 (66.7) | 4 (33.3) | 0.8 (0.2-2.5) | 2 (100) | 0 (0.0) | NA |
| **Education level** |  |  |  |  |  |  |
| Up to primary school |  |  |  | 203 (84.9) | 32 (15.1) | 2.4 (1.6-3.6) |
| Up to high school | NA | NA | NA | 1426 (88.2) | 190 (11.8) | 1.3 (1.4-2.4) |
| Higher education |  |  |  | 1432 (93.2) | 104 (6.8) | 1.0 (ref) |

**Table S3.**
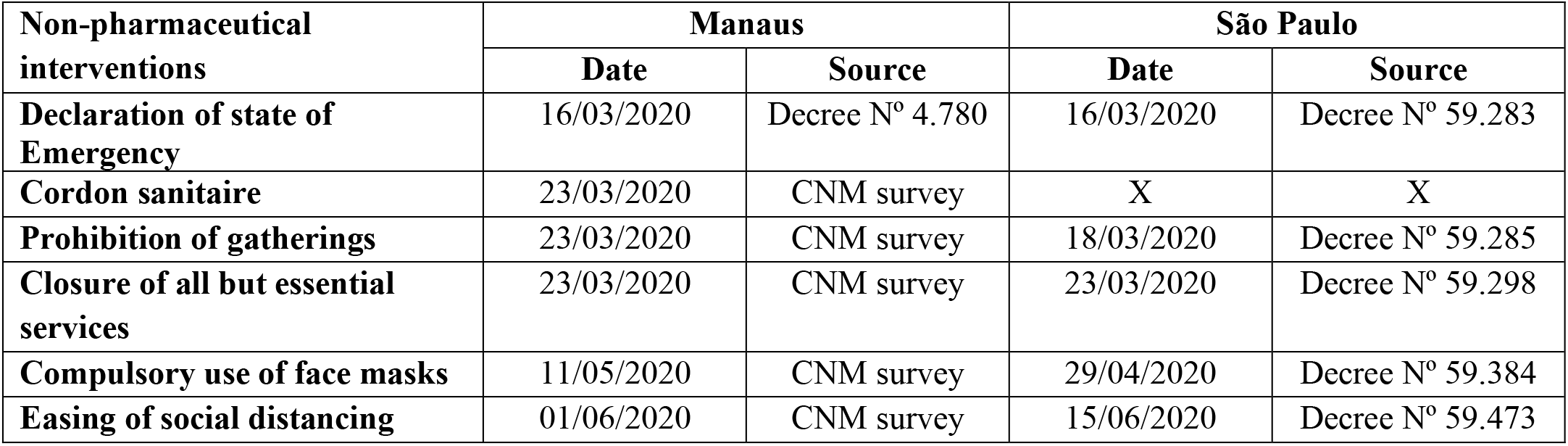
Implementation and easing of non-pharmaceutical measures in the municipalities of São Paulo and Manaus. Details on the CNM *(confederagao nacional de municipios)* survey can be found at (https://www.cnm.org.br/cms/biblioteca). Municipal-level decrees can be found at https://leismunicipais.com.br.

| Non-pharmaceutical interventions | Manaus |  | São Paulo |  |
| --- | --- | --- | --- | --- |
|  | Date | Source | Date | Source |
| **Declaration of state of Emergency** | 16/03/2020 | Decree N° 4.780 | 16/03/2020 | Decree N° 59.283 |
| **Cordon sanitaire** | 23/03/2020 | CNM survey | X | X |
| **Prohibition of gatherings** | 23/03/2020 | CNM survey | 18/03/2020 | Decree N° 59.285 |
| **Closure of all but essential services** | 23/03/2020 | CNM survey | 23/03/2020 | Decree N° 59.298 |
| **Compulsory use of face masks** | 11/05/2020 | CNM survey | 29/04/2020 | Decree N° 59.384 |
| **Easing of social distancing** | 01/06/2020 | CNM survey | 15/06/2020 | Decree N° 59.473 |

**Fig. S1.**
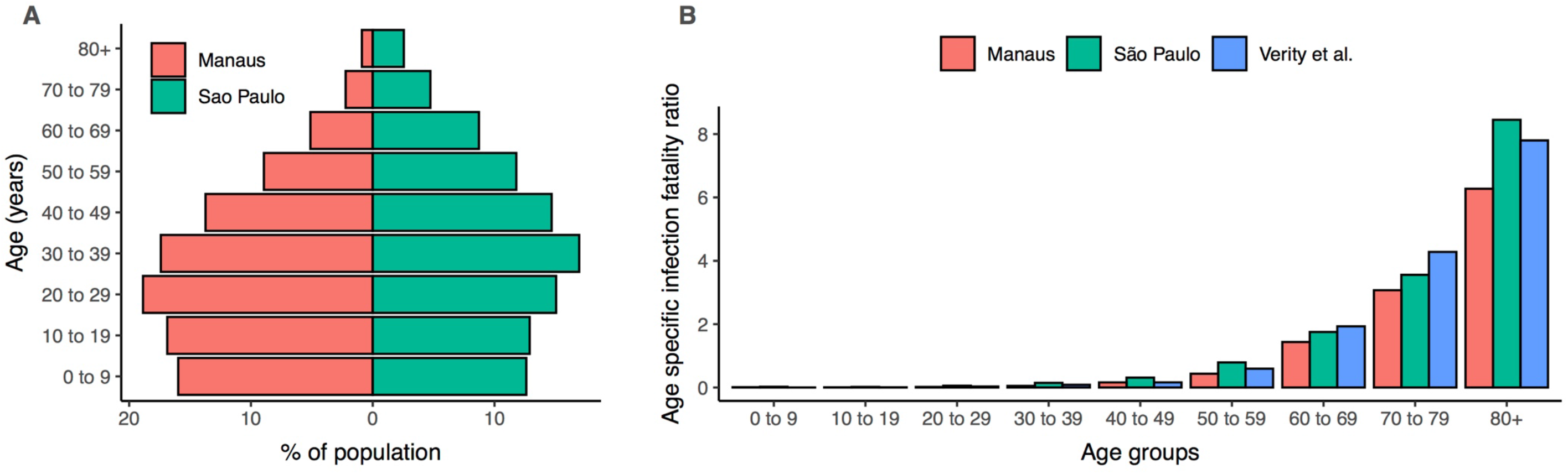
Age distribution in Manaus and São Paulo (A) and age-specific infection fatality ratios (%) for Manaus, São Paulo and from Verity et al^8^ (B). Age-specific IFRs were calculated using the seroprevalence in June, before significant seroreversion had occurred, and assuming equal prevalence across age groups. Deaths were taken from the SIVEP-Gripe database (https://covid.saude.gov.br/). Population data was taken from https://demografiaufrn.net/laboratorios/lepp/.

**Fig. S2.**
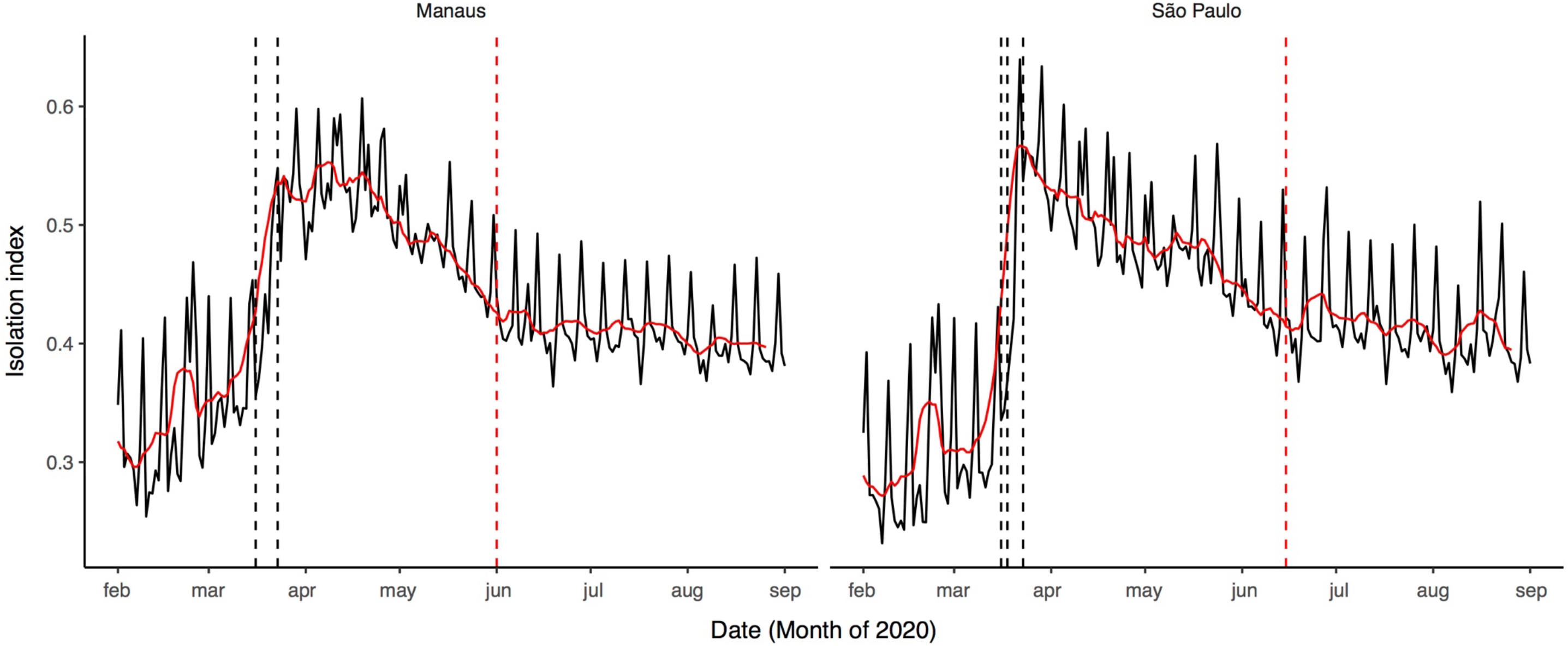
Isolation index calculated from cell phone data (https://mapabrasileirodacovid.inloco.com.br/pt/) for São Paulo and Manaus. Higher values for the isolation index indicate greater evidence of physical distancing. Red line represents the 7-day rolling average of the isolation index. Vertical black dashed lines show the timing at which non-pharmaceutical interventions were instigated; Vertical red dashed line is the date of relaxation of social distancing requirements (also see Table S3).

**Fig. S3.**
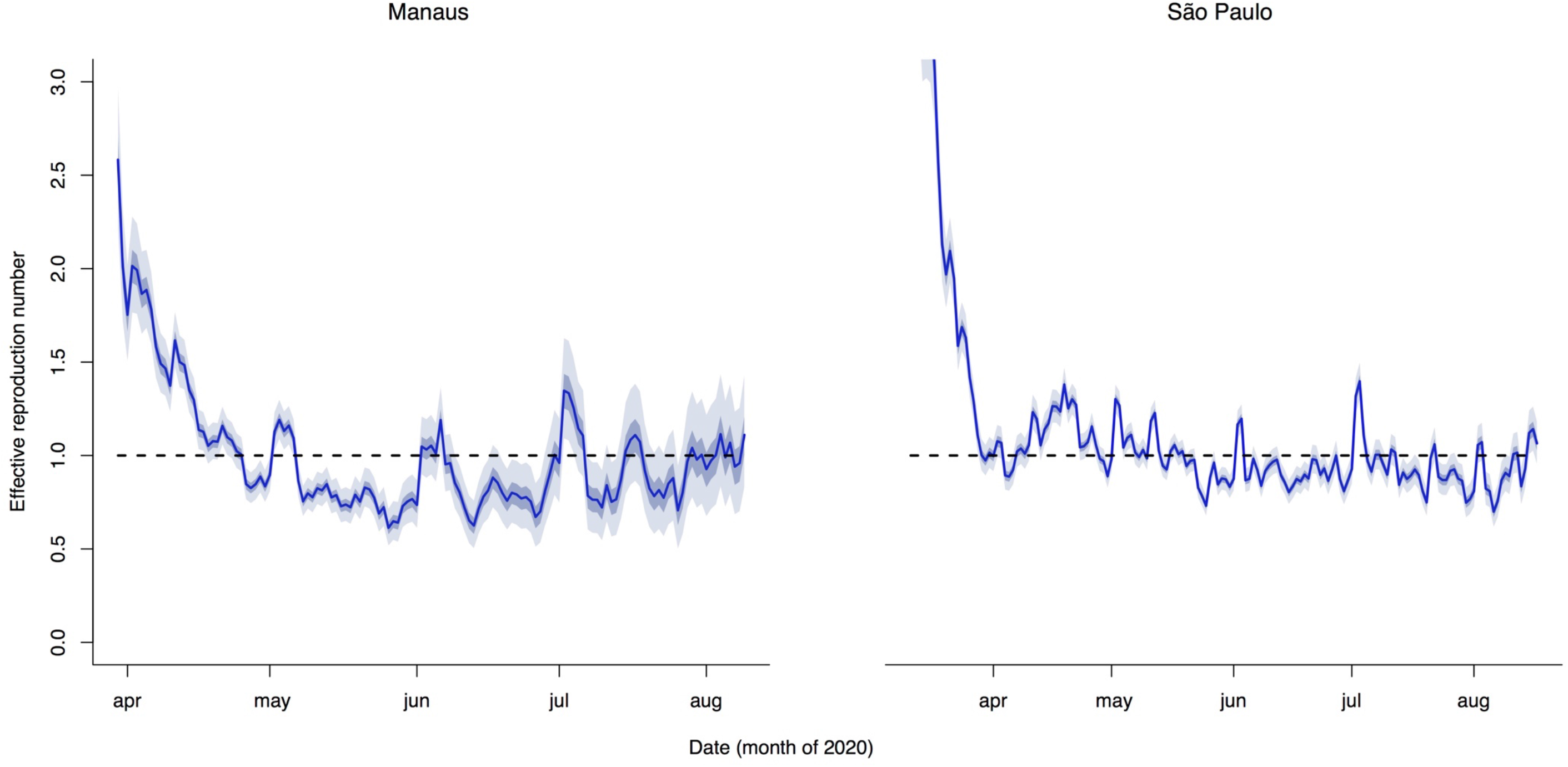
Effective reproduction number for Sâo Paulo and Manaus. Point estimate of the effective reproduction number is shown in dark blue and 95% confidence intervals in light blue.

**Fig. S4.**
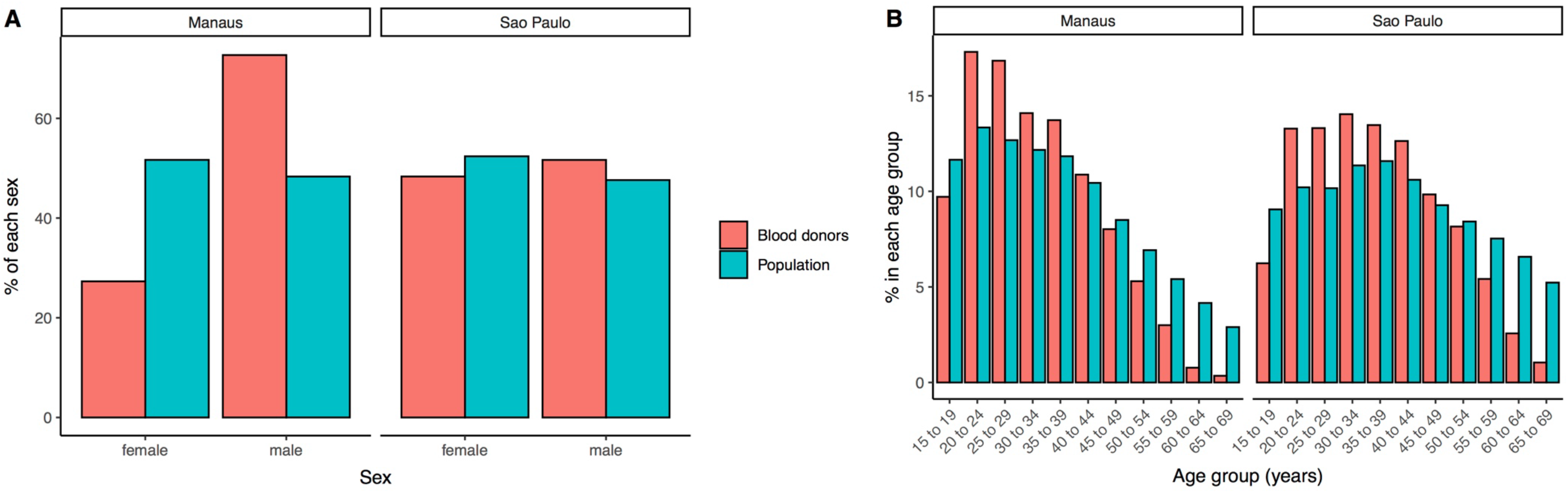
Comparison of the sex (A) and age (B) distributions of blood donors and the resident population in São Paulo and Manaus. The population percentages are out of the total population in the age range eligible to donate blood. Percentages for blood donors are out of all blood donations included in the study between February and August 2020. Population data was taken from https://demografiaufrn.net/laboratorio.

**Fig. S5.**
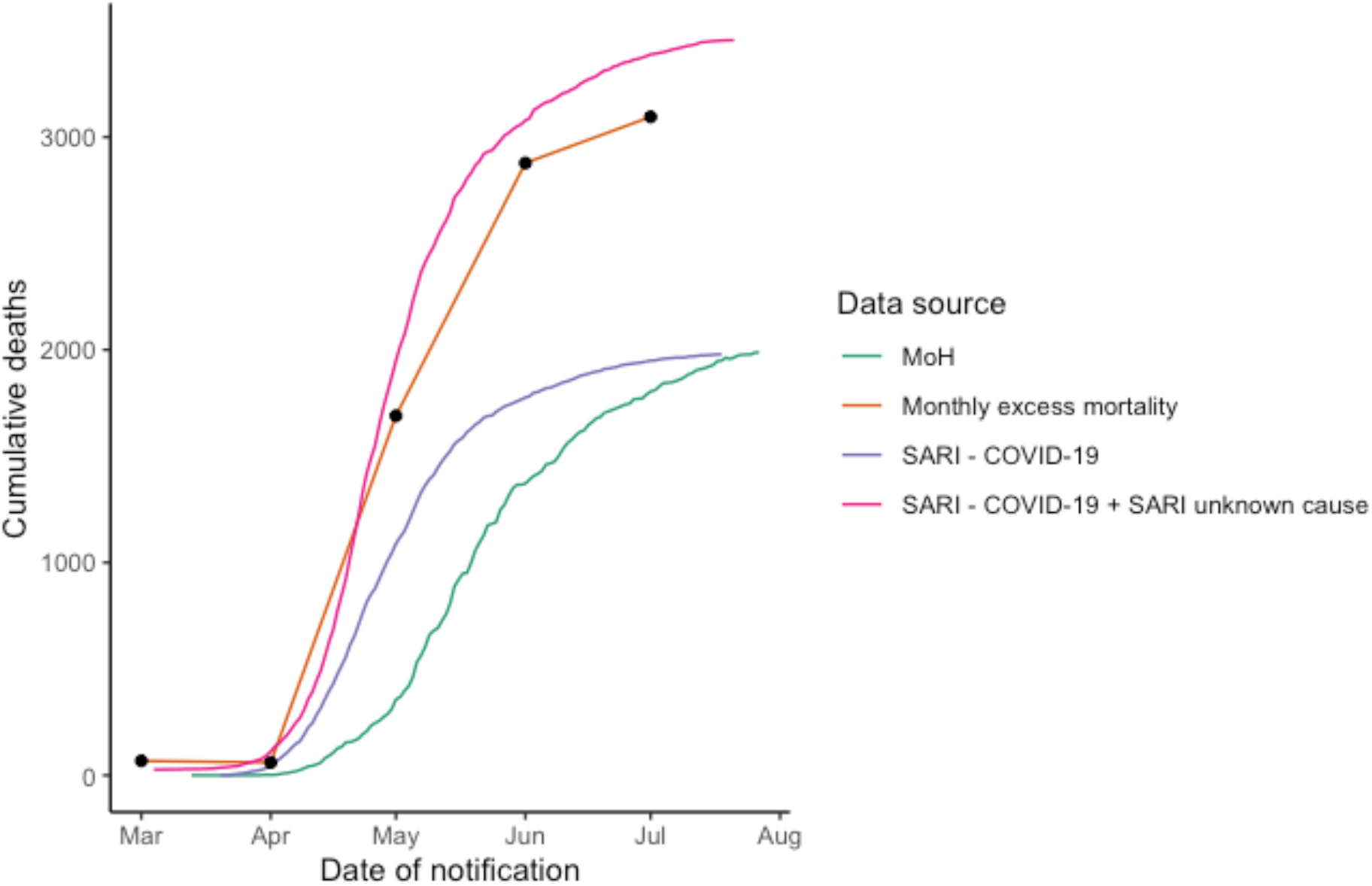
Cumulative deaths in Manaus according to multiple data sources. MoH – Ministry of Health official data source (https://covid.saude.gov.br/); excess mortality was calculated as the difference in total monthly deaths between 2020 and 2019 (data from https://transparencia.registrocivil.org.br/cartorios); Severe acute respiratory syndrome (SARI - SIVEP-Gripe https://opendatasus.saude.gov.br/dataset/bd-srag-2020).

**Fig. S6.**
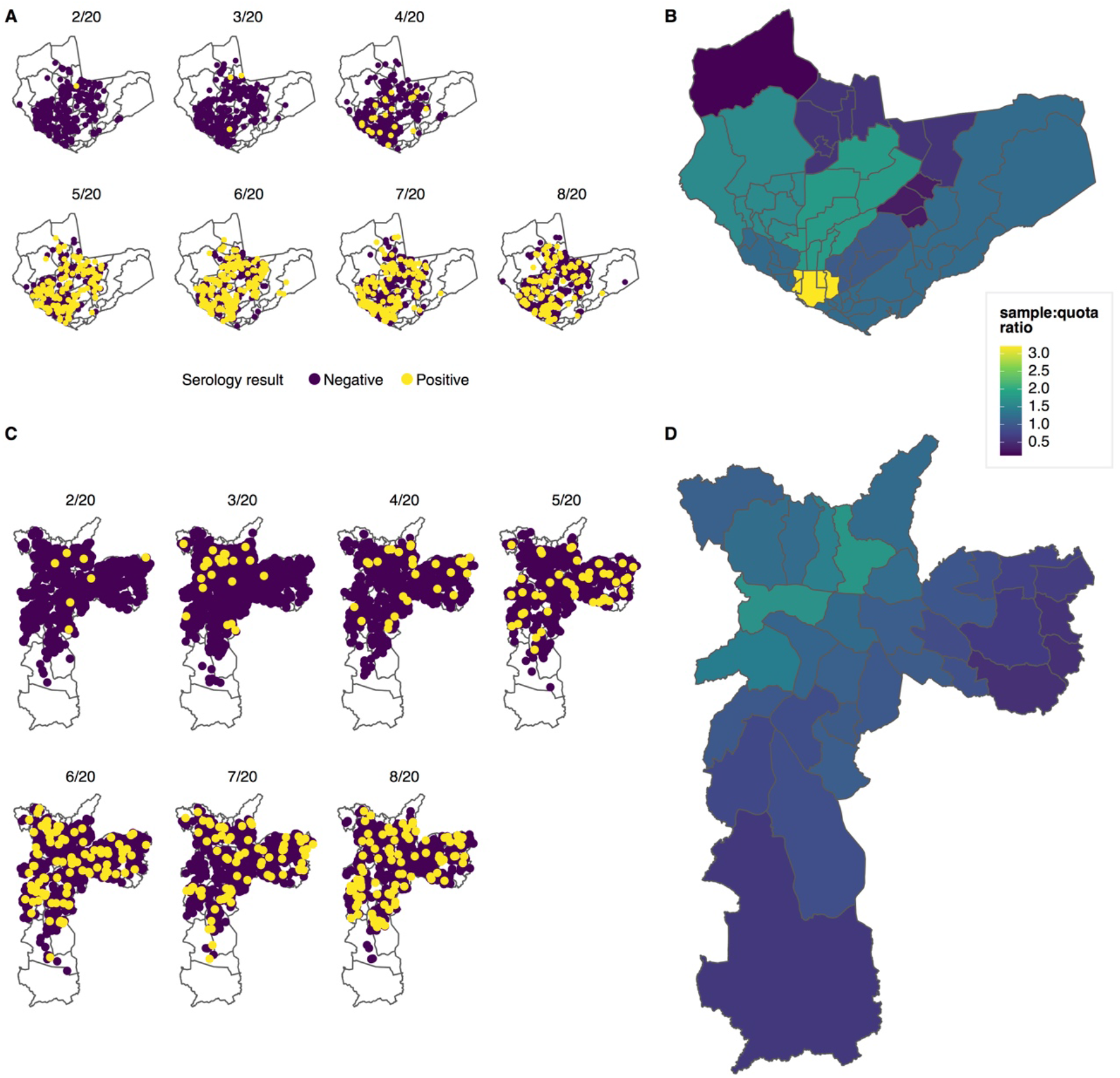
Spatial distribution of blood donors tested for SARS-CoV-2 antibodies in Manaus and São Paulo. Zip code location of blood donors tested in Manaus (A) and São Paulo (C) in each monthly sample between February and August 2020. In the case of Manaus 3,089/6,319 (48.9%) of tested samples had a recorded ZIP code and are shown on the figure. The degree to which the number of blood donation samples from each region of Manaus (B) and São Paulo (D) approximates the underlying population size is shown as the ratio between the number of samples tested and the sampling quota for each region after pooling data for February through August. Sampling quotas were re-scaled for Manaus to be proportional to a total of 3,089 with recorded ZIP. Lighter colors correspond to areas in which more blood samples were tested than indicated by the population size and dark colors where fewer were tested.

